# High Prevalence of Latent Tuberculosis Infection among Persons Deprived of Liberty in a Malaysian Prison

**DOI:** 10.1101/2024.10.19.24315804

**Authors:** Lu Zhang, Rumana Saifi, Adeeba Kamarulzaman, Ahsan Ahmad, Sangeeth Dhaliwal, Saidatul Hanida Mohd Yukhi, Nor Akma Ibrahim, Hui Moon Koh, Frederick L. Altice, Sheela V. Shenoi

**Author notes:** **Corresponding author:** Sheela V Shenoi, MD MPH, Yale University School of Medicine, 135 College Street, Suite 323, New Haven, CT, US 06510.

## Abstract

Rising tuberculosis incidence and mortality necessitate enhanced attention to prevention in neglected high-risk venues like correctional facilities. We sought to identify prevalence and correlates of latent TB infection (LTBI) in Malaysia’s largest prison. From October 2019 to January 2023, people deprived of liberty (PDL) entering Kajang Prison underwent tuberculin skin testing, sputum examination, chest X-ray, and blood tests for HIV, HCV, and C-reactive protein (CRP); PDL with active tuberculosis were excluded. Multivariable logistic regression identified independent correlates of LTBI. Among 601 men without tuberculosis, median age was 42 (IQR 36-50) years with high prevalences of HIV (8.8%) and HCV (43.4%). LTBI prevalence was 68.2% (95%CI[64.4%-71.8%]); independent risk factors included opioid use disorder (OUD, AOR=1.95; 95%CI[1.25-3.05]), pre-incarceration homelessness (AOR=1.89; 95%CI[1.13-2.50]), and HCV (AOR=1.68; 95%CI[1.13-2.50]). Pre-incarceration cannabis use (AOR=0.69; 95%CI[0.39-0.96]) was negatively associated with LTBI, which was also found in people with HIV (AOR=0.10; 95%CI[0.01-0.51]). Among people without HIV, having OUD (AOR=2.32; 95%CI[1.48-3.68]), HCV (AOR=1.64; 95%CI[1.09-2.48]) and CRP ≥5mg/ml (AOR=1.90; 95%CI[1.04-3.65]) were independently associated with LTBI, while methamphetamine use reduced odds of LTBI (AOR=0.58; 95%CI[0.33-0.98]). LTBI prevalence in Malaysia’s largest prison was high, highlighting the need for routine screening and implementation of TB preventive therapy in high-risk settings like prisons.

## Introduction

Tuberculosis (TB) remains a prominent global health concern despite extensive efforts to control its spread, especially in resource-limited correctional facilities in low- and middle-income country (LMIC) where conditions often contribute to the transmission of the disease.[1] The crowded inmate populations, increased vulnerability due to complicated comorbidities or medical histories, and continuous interaction increase the risk of spread.[2] Kajang’s prison for men, the largest in Malaysia, exemplifies these challenges.

Latent tuberculosis infection (LTBI) is a precursor for developing active TB disease, and can be prevented. [3] Coinfection with HIV complicates the LTBI landscape, making management even more challenging. Therefore, understanding LTBI prevalence and correlated factors, especially in high-risk populations such as in people deprived of liberty (PDL), is essential for devising effective preventive and therapeutic measures.[4]

Socio-demographic characteristics, substance use patterns, and other comorbidities have been identified as key determinants influencing risk for TB infection,[1,5,6] while substance use, particularly opioid dependence, has been correlated with an increased risk of acquiring TB.[7] Few studies have examined LTBI in the incarceration environment where the prevalence of opioid use disorder (OUD) and HIV are high. We therefore conducted a cross-sectional study of newly entering people to a prison to estimate the prevalence of LTBI, to discern potential correlates of LTBI and identify opportunities for better TB control.

## Methods

We conducted a cross-sectional study among PDL in Kajang prison, Malaysia’s largest prison for men, where HIV testing is mandatory and people with HIV (PWH) are segregated in dedicated housing units. Participants were eligible if within 2 days of prison entry, age ≥18 years old, held Malaysian citizenship, their prison sentence was >3 months, and no active TB in the past 2 years. Participants underwent a detailed interview combined with TB screening and diagnostic testing, including a battery of tuberculin skin testing (TST), sputum smear for acid-fast bacillus (AFB), sputum testing using Gene Xpert MTB/RIF, culture for Mycobacterium tuberculosis (MTB) (BACTEC MGIT 960 System) and chest X-ray (CXR). Those meeting criteria for TB, including positive findngs for TB culture, GeneXpert, and AFB smear were excluded. This study is reported following the STROBE recommendations (S1 Table) and was approved by the Institutional Review Boards of Yale University (IRB Number: 2000020053) and the University of Malaya (IRB Number: 201696-4220), and approved by the Malaysian Ministry of Health, and Prisons Department. Written informed consent was obtained from all participants prior to enrollment in the study.

Data were collected consecutively between 22^nd^ October 2019 and 19^th^ January 2023. After written informed consent, eligible participants underwent all TB testing requirements plus point-of-care HIV, phlebotomy for CRP, hepatitis C virus (HCV) and hepatitis B virus (HBV) antibody test. All positive HIV tests were confirmed using ELISA followed by CD4 and viral load testing and referral for antiretroviral therapy. In the structured survey, sociodemographic variables, substance use history, TB-associated medical histories including Bacille Calmette-Guerin (BCG) vaccine, TB contact, and symptom screen were collected. Pre-incarceration opioid dependence was defined using the Rapid Opioid Dependence Screen (RODS). CXR were performed at the prison by a mobile service within 1-2 weeks of entry, depending on COVID-19 restrictions, and reviewed by two radiologists; a third reviewer resolved any discordant results to assess for pulmonary TB. LTBI was confirmed when active TB was ruled out and the TST size ≥5mm for PWH and ≥10mm for PDL without HIV. The study was nested within a parent trial evaluating two regimens for TB preventive therapy. The sample size was influenced by the recruitment for the clinical trial of individuals determined to have no active TB disease. Initially, the trial enrolled only those with OUD, but on July 18, 2022, OUD was removed from the eligibility criteria to ensure generalizability.

REDCap was used to record the data and R (version 4.1.0) was used for statistical analysis. Descriptive statistics including counts and proportions were used to characterize categorical variables, while means with standard deviation (SD), medians with interquartile range (IQR), and 95% confidence intervals (CI) were used to characterize continuous variables. The primary outcome was the diagnosis of LTBI, operationalized as meeting TST criteria and having active TB excluded. Normality was checked among all continuous variables by QQ plot. Student’s t-test and the Wilcoxon rank sum test were used to compare parametric and non-parametric continuous variables, respectively. Chi-square test and Fisher’s exact test were used to compare categorical variables. All predictors with p-value ≤0.10 from the bivariate analysis were included into the multivariable model except for ‘history of heroin use’ due to its collinearity with diagnosis of pre-incarceration opioid dependence, confirmed using the RODS. Multivariable logistic regression was performed to identify independent correlates of LTBI using stepwise backward regression. Multicollinearity was assessed using the variance inflation factor (VIF).

## Results

Among the 935 PDL consecutively entering prison and meeting eligibility criteria, 926 (99.0%) participants enrolled. Among these, 80 were excluded as they had confirmed pulmonary TB. Another 245 were excluded due to missing data (Fig 1). The final analysis included 601 male participants with complete data. Table 1 provides the characteristics of the sample with the median age as 42 years (IQR 36-50) and 69.9% identified as Malay. Most participants were Muslim (73.7%), single (56.2%), and completed secondary school (75.4%). For the prior 6 months before entering prison, 85.5% of participants were employed full-time and their median monthly income was 324.3 US dollars, which is approximately 45% below the Malaysia 2022 poverty line of 594.4.[8] All participants had been previously incarcerated (median=8; IQR 4-14), with a median previous incarceration duration of 5.0 (IQR 2.7-10.0) years. Among participants, HIV prevalence was high (N=53, 8.8%) as was HCV infection (N=261, 43.4%). Only 6.3% had been hospitalized in the 6 months prior to entering prison. In total, 477 (79.4%) participants had OUD and 253 (42.1%) participants reported injecting drugs before being incarcerated. For participants included after July 18, 2022, the prevalence of OUD was 51.9% (134/258). The prevalence of illegal drug use was high: heroin ranked highest (509 [84.7%]), followed by methamphetamine (496 [82.5%]), and marijuana (428 [71.2%]).

**Table 1.** Comparison of full baseline characteristics, stratified by LTBI status (N=601)

| Predictors (Mean [SD] / N [%]) | LTBI-negative<br>(N=191) | LTBI-positive<br>(N=410) | Total (N=601) | P value<sup>1</sup> |
| --- | --- | --- | --- | --- |
| Age | 41.4 (10.0) | 43.34 (9.6) | 42.7 (9.8) | 0.025* |
| Race |  |  |  | 0.564 |
| Malay | 136 (71.2%) | 284 (69.3%) | 420 (69.9%) |  |
| Chinese | 18 (9.4%) | 38 (9.3%) | 56 (9.3%) |  |
| Indian | 30 (15.7%) | 79 (19.3%) | 109 (18.1%) |  |
| Other | 7 (3.7%) | 9 (2.2%) | 16 (2.7%) |  |
| Religion |  |  |  | 0.379 |
| Islam | 147 (77.0%) | 296 (72.2%) | 443 (73.7%) |  |
| Christianity | 12 (6.3%) | 20 (4.9%) | 32 (5.3%) |  |
| Buddhism | 11 (5.8%) | 27 (6.6%) | 38 (6.3%) |  |
| Hinduism | 19 (9.9%) | 64 (15.6%) | 83 (13.8%) |  |
| Other | 2 (1.0%) | 3 (0.7%) | 5 (0.8%) |  |
| Marital status |  |  |  | 0.391 |
| Single | 102 (53.4%) | 236 (57.6%) | 338 (56.2%) |  |
| Married | 41 (21.5%) | 87 (21.2%) | 128 (21.3%) |  |
| Separated | 10 (5.2%) | 27 (6.6%) | 37 (6.2%) |  |
| Widowed | 38 (19.9%) | 60 (14.6%) | 98 (16.3%) |  |
| Education level |  |  |  | 0.351 |
| Primary school and below | 35 (18.3%) | 95 (23.2%) | 130 (21.6%) |  |
| Secondary school | 149 (78.0%) | 304 (74.1%) | 453 (75.4%) |  |
| Above Secondary school | 7 (3.7%) | 11 (2.7%) | 18 (3.0%) |  |
| Employed prior to incarceration | 171 (89.5%) | 343 (83.7%) | 514 (85.5%) | 0.057 |
| Income prior to incarceration<br>(USD/month) | 410.7(512.3) | 450.9 (1690.5) | 438.2 (1425.3) | 0.748 |
| Hospitalization in the past 6<br>months | 12 (6.3%) | 26 (6.3%) | 38 (6.3%) | 0.978 |
| Number of prior incarcerations | 9.0 (7.3) | 10.5 (7.6) | 10.0 (7.6) | 0.030* |
| IVDU | 60 (31.4%) | 193 (47.1%) | 253 (42.1%) | < 0.001*** |
| Opioid dependence | 130 (68.1%) | 347 (84.6%) | 477 (79.4%) | < 0.001*** |
| BCG vaccine | 183 (95.8%) | 399 (97.3%) | 582 (96.8%) | 0.326 |
| Homelessness | 17 (8.9%) | 76 (18.5%) | 93 (15.5%) | 0.002** |
| TB contact | 30 (15.7%) | 93 (22.7%) | 123 (20.5%) | 0.048* |
| Marijuana | 152 (79.6%) | 276 (67.3%) | 428 (71.2%) | 0.002** |
| Heroin | 149 (78.0%) | 360 (87.8%) | 509 (84.7%) | 0.002** |
| Cocaine | 21 (11.0%) | 50 (12.2%) | 71 (11.8%) | 0.671 |
| Methamphetamine | 168 (88.0%) | 328 (80.0%) | 496 (82.5%) | 0.017* |
| MDMA | 26 (13.6%) | 45 (11.0%) | 71 (11.8%) | 0.351 |
| GHB | 4 (2.1%) | 7 (1.7%) | 11 (1.8%) | 0.742 |
| Ketamine | 48 (25.1%) | 84 (20.5%) | 132 (22.0%) | 0.200 |
| Inhalants | 23 (12.0%) | 61 (14.9%) | 84 (14.0%) | 0.350 |
| Kratom leaves | 70 (36.6%) | 134 (32.7%) | 204 (33.9%) | 0.339 |
| BMI | 22.1 (3.2) | 22.1 (2.9) | 22.1 (3.0) | 0.730 |
| HIV | 15 (7.9%) | 38 (9.3%) | 53 (8.8%) | 0.569 |
| Currently taking ART | 8 (4.2%) | 28 (6.8%) | 36 (6.0%) | 0.184 |
| HIV Viral load ≥200 copies/mL | 11 (5.8%) | 20 (4.9%) | 31 (5.2%) | 0.649 |
| HCV | 58 (30.4%) | 203 (49.5%) | 261 (43.4%) | < 0.001*** |
| HBV | 1 (0.5%) | 6 (1.5%) | 7 (1.2%) | 0.317 |
| CRP ≥5mg/L | 18 (9.4%) | 58 (14.1%) | 76 (12.6%) | 0.105 |
| CXR concerning for TB | 36 (18.8%) | 121 (29.5%) | 157 (26.1%) | 0.006** |

Abbreviations: LTBI, latent tuberculosis infection; SD, standard deviation; IVDU, intravenous drug user; BCG, bacille Calmette-Guerin; TB, tuberculosis; MDMA, 3,4-Methylenedioxymethamphetamine; GHB, Gamma-hydroxybutyrate; BMI, Body mass index; HIV, human immunodeficiency virus; ART, antiretroviral therapy; HCV, hepatitis C virus; HBV, hepatitis B virus; CRP, C-reactive protein; CXR, chest X-ray.
1: \*P<0.05; \*\*P<0.01; \*\*\*P<0.001

**Fig 1.**
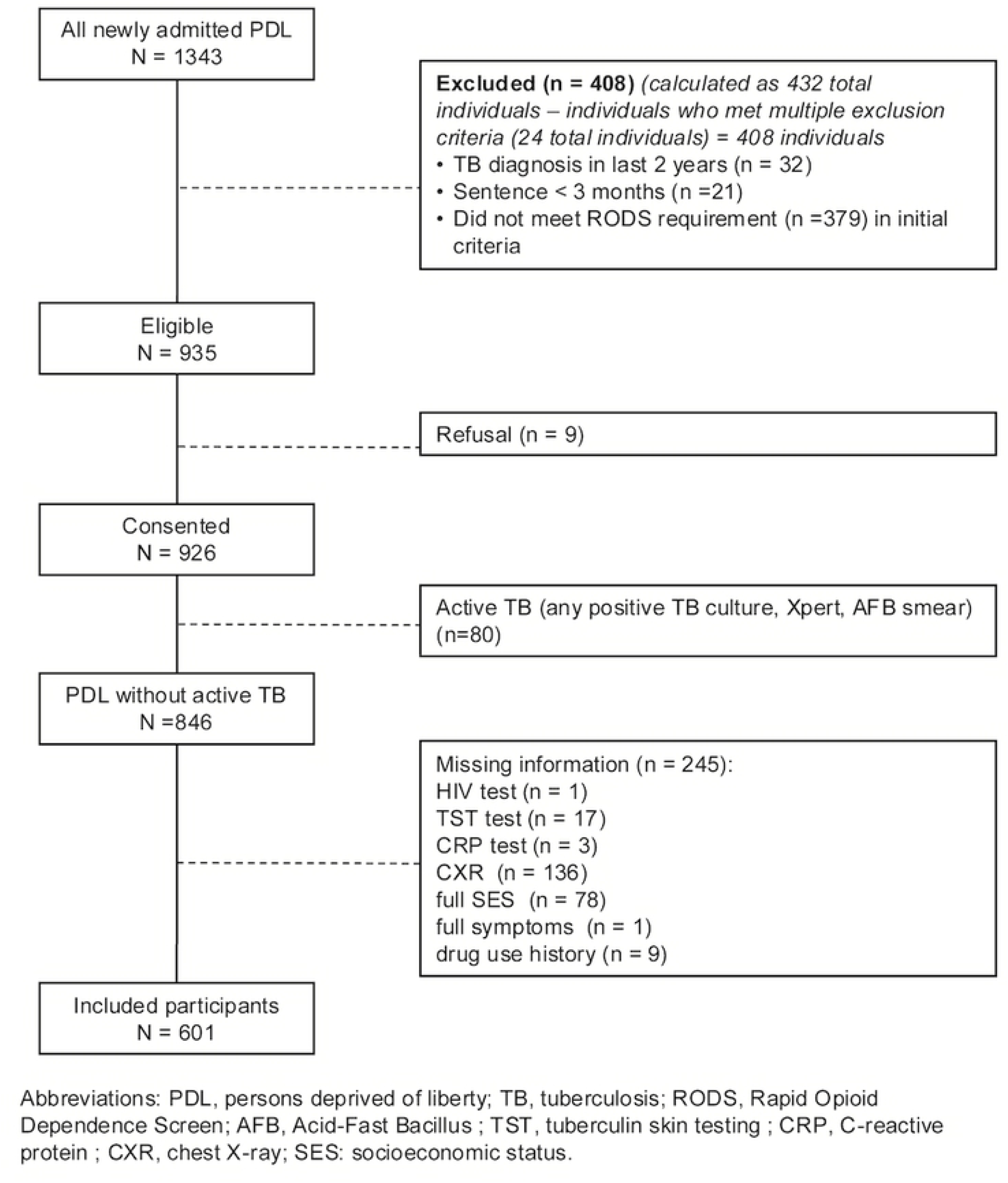
Patient flow chart.

The prevalence of LTBI was 68.2% (95%CI: 64.4%-71.8%) and higher among PWH (77.4%; 95%CI: 67.2%-87.7%).The LTBI prevalence among PDL without HIV was 67.9% (95%CI: 64.0%-71.8%). The vast majority (96.8%) of participants reported receiving the BCG vaccine, 123 (20.5%) reported contact with someone with TB, and 15.5% reported a history of homelessness (Table 1).

On multivariable analysis, independent risk factors for LTBI among all PDL (Table 2) included OUD history (Adjusted odds ratio [AOR] = 1.95 [95%CI: 1.25-3.05], p=0.003), history of homelessness (AOR = 1.89[95%CI: 1.09-3.45], p=0.03) and having hepatitis C (AOR = 1.68 [95%CI: 1.13-2.50], p=0.01). Marijuana use was the only independent factor associated with less risk of LTBI (AOR = 0.69 [95%CI: 0.39-0.96], p=0.035).

**Table 2.** Correlates of LTBI among new entrants to prison (N=601)

| Variables<sup>1</sup> | Univariable analysis |  | Multivariable analysis<sup>2</sup> |  |
| --- | --- | --- | --- | --- |
|  | OR (95% CI) | P value | AOR (95% CI) | P value<sup>3</sup> |
| Age | 1.021<br>(1.003-1.039) | 0.026* | -- | -- |
| Pre-incarceration: |  |  |  |  |
| Employed prior to incarceration | 0.599<br>(0.344-1.002) | 0.059 | -- | -- |
| Number of prior incarcerations | 1.027<br>(1.003-1.053) | 0.031* | -- | -- |
| Drug injection | 1.942<br>(1.357-2.801) | <0.001*** | -- | -- |
| Opioid dependence | 2.584 | <0.001*** | 1.952 | 0.003** |
|  | (1.722-3.882) |  | (1.252-3.047) |  |
| Homelessness | 2.329 | 0.003** | 1.891 | 0.030* |
|  | (1.365-4.186) |  | (1.087-3.452) |  |
| TB contact | 1.574 | 0.050 | -- | -- |
|  | (1.011-2.510) |  |  |  |
| Marijuana | 0.528 | 0.002** | 0.619 | 0.035* |
|  | (0.348-0.789) |  | (0.393-0.960) |  |
| Methamphetamine | 0.548 | 0.018* | 0.628 | 0.098 |
|  | (0.326-0.888) |  | 0.356-1.077) |  |
| Heroin | 2.030 | 0.002** | -- | -- |
|  | (1.287-3.189) |  |  |  |
| After-incarceration: |  |  |  |  |
| HCV | 2.249 | <0.001*** | 1.677 | 0.011* |
|  | (1.569-3.253) |  | (1.130-2.501) |  |
| CXR concerning for TB | 1.803 | 0.006** | 1.440 | 0.104 |
|  | (1.194-2.773) |  | (0.934-2.255) |  |

Abbreviations: LTBI, latent tuberculosis infection; OR, odds ratio; AOR: adjusted odds ratio; CI, confidence interval; IVDU, intravenous drug user; HCV, hepatitis C; CRP: C-reactive protein; CXR, chest X-ray; TB, tuberculosis.
1: Only variables with P values <0.10 in univariable analysis are listed here.
2: Stepwise backward regression was applied in multivariable model. No multicollinearity detected since all three adjusted models had VIFs less than 1.2.
3: \*P<0.05; \*\*P<0.01; \*\*\*P<0.001

For PWH (Table 3), comorbid HCV increased the odds of LTBI (AOR = 3.86 [95%CI 0.88-18.6], p=0.078) while the history of marijuana use was associated with lower odds for LTBI (AOR = 0.10 [95%CI: 0.01-0.51], p=0.01). For PDL without HIV (Table 4), participants who had OUD history (AOR = 2.32 [95%CI: 1.48-3.68], p<0.001), having hepatitis C (AOR = 1.64 [95%CI: 1.09-2.48], p=0.018) and CRP ≥5mg/ml (AOR = 1.90 [95%CI: 1.04-3.65], p=0.044) are independent risk factors for LTBI, while the history of methamphetamine use was associated with lower odds of LTBI (AOR = 0.58 [95%CI: 0.33-0.98], p=0.048). All PWH had a history of homelessness or hospitalization in the past 6 months, which indicates that these two factors may be potential LTBI risk factors among PWH, which need more data to explore.

**Table 3.**
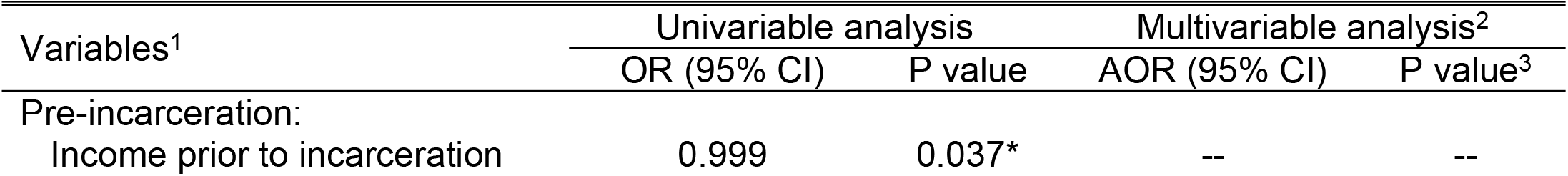
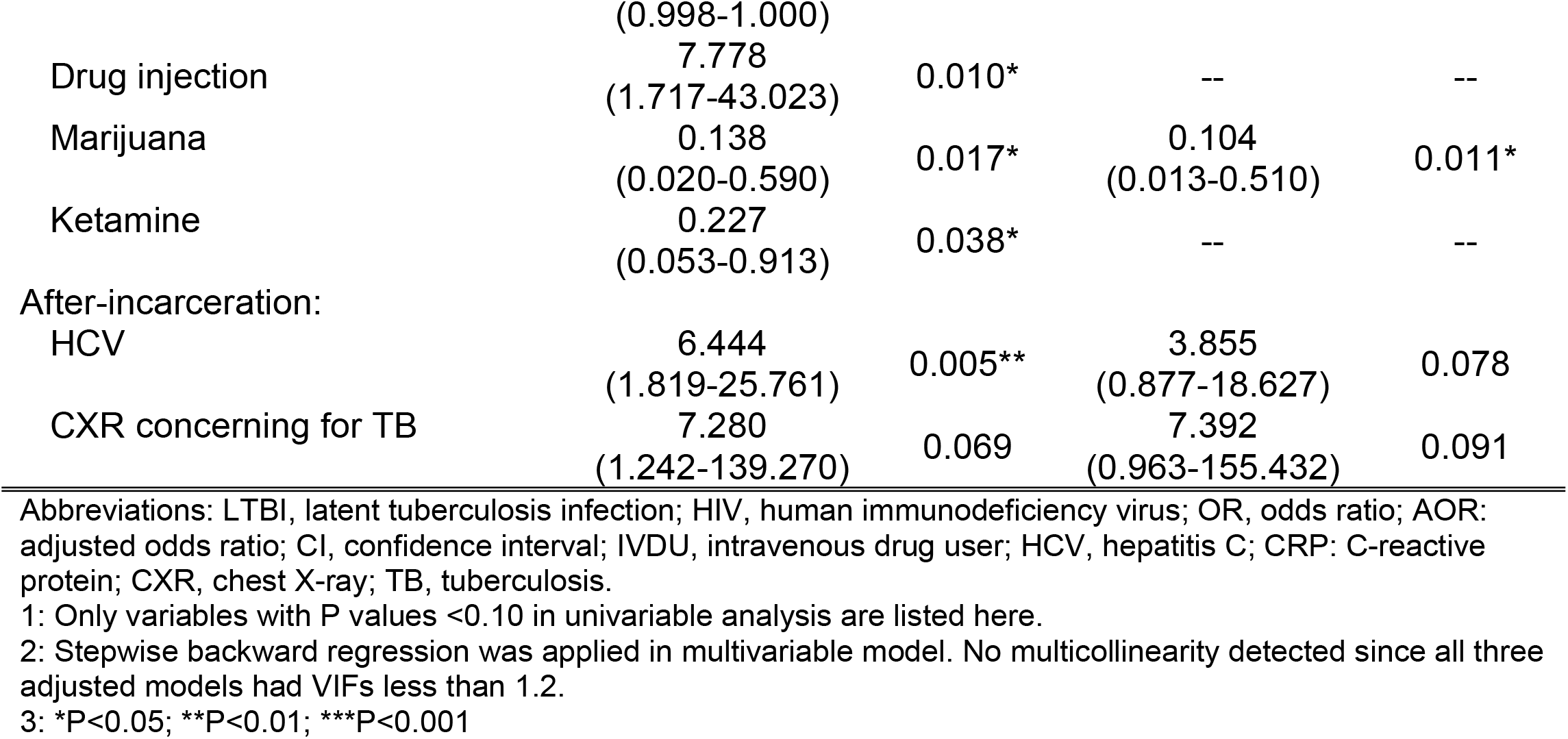
Correlates of LTBI among people with HIV (N=53)

| Variables<sup>1</sup> | Univariable analysis |  | Multivariable analysis<sup>2</sup> |  |
| --- | --- | --- | --- | --- |
|  | OR (95% CI) | P value | AOR (95% CI) | P value<sup>3</sup> |
| Pre-incarceration: |  |  |  |  |
| Income prior to incarceration | 0.999 | 0.037* | -- | -- |
|  | (0.998-1.000) |  |  |  |
| Drug injection | 7.778<br>(1.717-43.023) | 0.010* | -- | -- |
| Marijuana | 0.138<br>(0.020-0.590) | 0.017* | 0.104<br>(0.013-0.510) | 0.011* |
| Ketamine | 0.227<br>(0.053-0.913) | 0.038* | -- | -- |
| After-incarceration: |  |  |  |  |
| HCV | 6.444<br>(1.819-25.761) | 0.005** | 3.855<br>(0.877-18.627) | 0.078 |
| CXR concerning for TB | 7.280<br>(1.242-139.270) | 0.069 | 7.392<br>(0.963-155.432) | 0.091 |

Abbreviations: LTBI, latent tuberculosis infection; HIV, human immunodeficiency virus; OR, odds ratio; AOR: adjusted odds ratio; CI, confidence interval; IVDU, intravenous drug user; HCV, hepatitis C; CRP: C-reactive protein; CXR, chest X-ray; TB, tuberculosis.
1: Only variables with P values <0.10 in univariable analysis are listed here.
2: Stepwise backward regression was applied in multivariable model. No multicollinearity detected since all three adjusted models had VIFs less than 1.2.
3: \*P<0.05; \*\*P<0.01; \*\*\*P<0.001

**Table 4.**
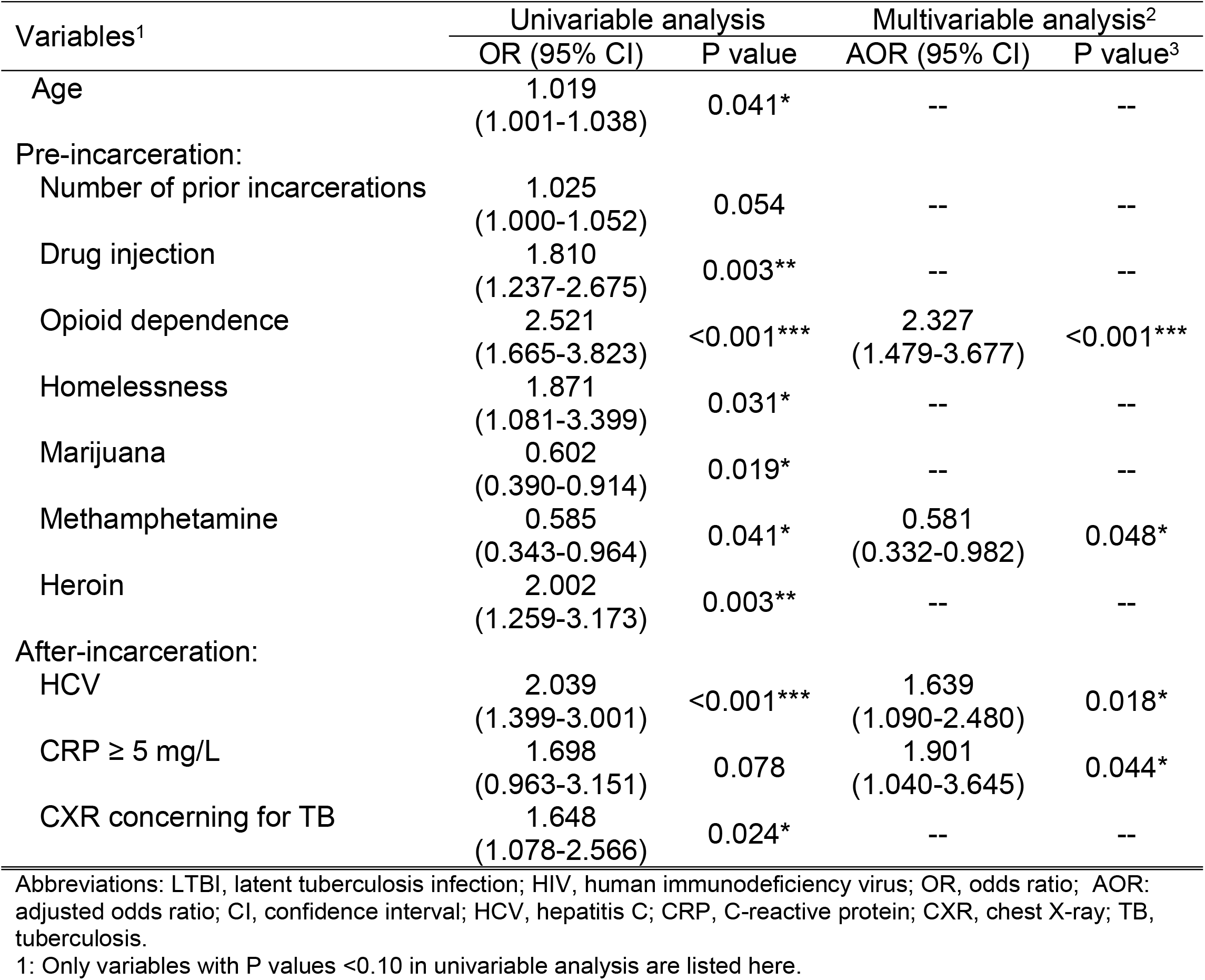
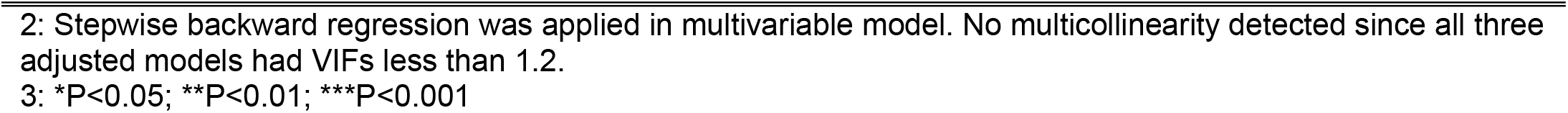
Correlates of LTBI among people without HIV (N=548)

| Variables<sup>1</sup> | Univariable analysis |  | Multivariable analysis<sup>2</sup> |  |
| --- | --- | --- | --- | --- |
|  | OR (95% CI) | P value | AOR (95% CI) | P value<sup>3</sup> |
| Age | 1.019<br>(1.001-1.038) | 0.041* | -- | -- |
| Pre-incarceration: |  |  |  |  |
| Number of prior incarcerations | 1.025<br>(1.000-1.052) | 0.054 | -- | -- |
| Drug injection | 1.810<br>(1.237-2.675) | 0.003** | -- | -- |
| Opioid dependence | 2.521<br>(1.665-3.823) | <0.001*** | 2.327<br>(1.479-3.677) | <0.001*** |
| Homelessness | 1.871<br>(1.081-3.399) | 0.031* | -- | -- |
| Marijuana | 0.602<br>(0.390-0.914) | 0.019* | -- | -- |
| Methamphetamine | 0.585<br>(0.343-0.964) | 0.041* | 0.581<br>(0.332-0.982) | 0.048* |
| Heroin | 2.002<br>(1.259-3.173) | 0.003** | -- | -- |
| After-incarceration: |  |  |  |  |
| HCV | 2.039<br>(1.399-3.001) | <0.001*** | 1.639<br>(1.090-2.480) | 0.018* |
| CRP ≥ 5 mg/L | 1.698<br>(0.963-3.151) | 0.078 | 1.901<br>(1.040-3.645) | 0.044* |
| CXR concerning for TB | 1.648<br>(1.078-2.566) | 0.024* | -- | -- |

Abbreviations: LTBI, latent tuberculosis infection; HIV, human immunodeficiency virus; OR, odds ratio; AOR: adjusted odds ratio; CI, confidence interval; HCV, hepatitis C; CRP, C-reactive protein; CXR, chest X-ray; TB, tuberculosis.
1: Only variables with P values <0.10 in univariable analysis are listed here.
2: Stepwise backward regression was applied in multivariable model. No multicollinearity detected since all three adjusted models had VIFs less than 1.2. 3: \*P<0.05; \*\*P<0.01; \*\*\*P<0.001

## Discussion

To our knowledge, this is the first systematic assessment of LTBI prevalence using microbiological assessment to gauge the absence of active TB among PDL in Malaysia, a upper middle-income country with a high prevalence of HIV, TB, HCV, and OUD.[9,10] In this large cross-sectional study assessing LTBI among PDL, we found a high LTBI prevalence at 68.2%. Among PDL overall, OUD, a history of homelessness, and hepatitis C were associated with higher odds of LTBI. No specific LTBI risk factors were identified in the subset of PDL with HIV. This high LTBI prevalence highlights the need for targeted screening and preventive therapy in correctional facilities.

Malaysia is a medium TB burden country with a stable LTBI prevalence rate from 28.2% in 1990 to 25.5% in 2019 among the general population,[11] much lower than among PDL from our study. Prisons are known to be hotspots for TB transmission given the vulnerabilities of PDL and the structural characteristics of prisons that facilitate transmission.[12,13] Data on the prevalence of LTBI in prisons is sparse. A study in western Ethiopia demonstrated a LTBI prevalence of 51.2%; a study in Columbia showed 46.4% prevalence among PDL with less than 3 months of incarceration but 70.5% prevalence in cases with more than 1 year of incarceration.[14,15] These values were higher than recent prison studies in high-income countries: 7.1% in the United Kingdom,[16] and 13.4% in Texas, the United States.[17] While the LTBI prevalence of another prison in Malaysia 10 years ago was reported as 87.6%, higher than our study result, though this study was based on symptom screen and TST only.[2] Rising prevalence of HIV, HCV, and PWUD are major contributors to these epidemics among incarcerated populations.[5,18] As increasing evidence identifies prisons as TB transmission hotspots and sources of active tuberculosis for surrounding communities,[19] there is increased interest in promoting TB preventive therapy in prison settings.[20] The WHO consolidated guidelines on TB prevention have recommended all patients living with HIV without active TB should receive TB preventive therapy, and in 2020, the WHO expanded guidance to recommend that people in prison receive systematic LTBI testing and treatment as well.[21] As prisons concentrate high-risk populations, PDL should be screened systematically at their admissions, with subsequent TB preventive therapy.[6,20] Further evidence about implementation models of TB preventive therapy in prison settings, and the impact of TB preventive therapy among PDL are needed.

Our study showed OUD and hepatitis C infection were both risk factors among general PDL and prisoners living without HIV, and OUD had higher ORs than HCV infection in both subsets. HCV is likely a marker of substance use, particularly intravenous drug users (IVDU), though IVDU was not significant on multivariable analysis among all three subsets in our study. This relationship has also been reported in other developed countries with a high prevalence of IVDU.[22,23] PWUD has been proved as a risk group of LTBI by many studies and WHO,[7,16,21,24,25] but there has been debate on whether this is due to behavior, i.e. close contact with someone with active TB disease or because of substance use-related immunologic susceptibility.[23] This study showed that neither intravenous access nor inhalants showed an independent association with LTBI under the adjusted regression model, and opioid use is the only drug-related risk factor. The lack of specific risk correlates among PDL with HIV may be due to a limited sample size. In previous studies, elevated CRP had shown value as a screening tool for active TB among PWH.[26] As for LTBI, CRP was not significantly correlated among PWH, but elevated CRP was an independent risk factor among PDL without HIV based on our analysis.

We collected substance use details among all PWUD and surprisingly found marijuana was significantly associated with less risk of LTBI among general PDL and among PDL with HIV, while methamphetamine showed lower odds of LTBI among PDL without HIV. Multiple drug use could lead to a lower frequency of opioid use even if the participant was identified with OUD. A systematic review found that marijuana use may be a risk factor for LTBI, though this was based on only 2 cross-sectional studies. It is plausible that non-opioid drug use is less risky for LTBI since there may be less individual close contact than with OUD.[27] Clinicians assessing patients for tuberculosis frequently lump together all substance use; further research could allow for further discrimination of risk.

We explored LTBI prevalence and respective correlates based on HIV status in correctional settings, which emphasizes the importance of screening and care pathways in different populations. It also highlights the importance of screening upon prison entry as early evaluation can identify high-risk groups for active TB and support timely active TB and LTBI treatment initiation and completion. Our findings identify substance use as a significant risk factor for LTBI highlighting the importance of substance use screening and treatment integration into TB care in prison settings. Additionally, the study’s use of comprehensive diagnostic methods (TB culture, Xpert, AFB smear) ensured a robust exclusion of active TB.

We recognize several limitations. In the parent study, investigators were denied access to the women’s prison, thus the findings identified in this evaluation may not be generalizable to women who are incarcerated. Next, the sample has a high prevalence of OUD since the parent study initially included OUD as an eligibility criterion, and thus the findings may not be generalizable to prison settings where OUD is less prevalent, though this scenario is uncommon based on available global data.[2,5,14,28] Furthermore, the study relied on TST, which is less sensitive and specific than interferon-gamma release assays (IGRA).[29] The inclusion of adults lowers the false-positive risk among BCG-vaccinated population since the effects of BCG wane within 8– 10 years[2]. Lastly, the proportion of PWH was small, limiting analyses for this subgroup.

In conclusion, the prevalence of LTBI in Malaysian prisoners was high, nearly three times of the general population, which highlights the need for screening and the implementation of TB preventive therapy to reduce risk amongst PDL, staff, and to the general population. Among newly admitted PDL, opioid use history and HCV are the main risk factors for LTBI, especially for PDL living without HIV. LTBI is also associated with the history of homelessness for general PDL and elevated CRP for PDL living without HIV. Substance use including marijuana and methamphetamine are associated with lower odds of LTBI. Hence, screening algorithms based on HIV status, HCV status, drug use history, and homelessness may be useful in stratifying risk upon prison entry. Further evidence-based approaches are also needed in the implementation of TB preventive therapy among PDL.

## Data Availability

All relevant data are within the manuscript and its Supporting Information files.

## Acknowledgment

We are grateful to Kajang Prison, Prisons Department, and the Malaysian Ministry of Health. We thank our staff and colleagues at University of Malaya Centre of Excellence for Research in AIDS (CERIA) and Yale University. Additionally, we convey our sincere thanks to the persons deprived of liberty who consented to partake in the survey. We have no conflicts of interest to declare.

## Supporting information

**S1 Table. STROBE Checklist of cross-sectional studies**

